# Power spectra density and similarity analysis of COVID-19 mortality waves across countries

**DOI:** 10.1101/2024.03.06.24303885

**Authors:** Elias Manjarrez, Erick F Delfin, Saul M Dominguez-Nicolas, Amira Flores

## Abstract

Johns Hopkins University CSSE documented waves of oscillatory COVID-19 mortality patterns worldwide during the COVID-19 pandemic. Here, we calculated the power spectrum density (PSD) of these COVID-19 mortality waves in 199 countries from January 22, 2020, to March 9, 2023. We identified two dominant peaks in the grand averaged PSD: one at a frequency of 1.15 waves per year (i.e., one wave every 10.4 months) and another at 2.7 waves per year (i.e., one wave every 4.4 months). Moreover, we performed a cosine similarity analysis of these PSD patterns among all the countries. The results showed a cosine similarity index distribution that was negatively skewed, with a skewness of −0.54 and a global median of cosine similarity index of 0.84, thus revealing a remarkable similarity in the dominant peaks of the COVID-19 mortality waves. These findings could be helpful if a future pandemic of a similar scale occurs so that effective confinement measures or other actions could be planned during these two identified periods, ensuring more assertive public health policies in advance.

## Introduction

Although the Spanish flu occurred over 100 years ago (Berche, 2022; Lista et al., 2023; Doran et al., 2024), no nation was prepared for a pandemic of this scale in modern times. This lack of preparation and the variety of transportation systems between nations contributed to the COVID-19 pandemic spreading rapidly and affecting all countries worldwide. This, in turn, caused a total collapse in healthcare systems in many countries from the onset of the disease (Dong et al., 2022). Another contributing factor was the high virulence of the SARS-CoV-2 virus, which, combined with the massive global population, facilitated a dramatic increase in infections from the start, leading to a high rate of infections in many countries within weeks (Hass and Arsanjani, 2021). Hence, the COVID-19 pandemic resulted in over 7 million deaths worldwide. In this context, the number of fatalities proves that the pandemic control was unsuccessful without vaccination. Therefore, a systematic analysis of COVID-19 mortality patterns worldwide would be helpful for future generations if a new pandemic of a similar scale could occur.

Fortunately, it is now possible to obtain data on the daily number of SARS-CoV-2 infections, active cases, and COVID-19 deaths worldwide thanks to databases such as "Worldometer" and Johns Hopkins University CSSE (Dong et al., 2022). These databases were created to facilitate future studies on the factors contributing to the virus’s spread. For this reason, infections and confirmed COVID-19 deaths have been geographically mapped in space and time (Hass and Arsanjani, 2021; Dong et al., 2022). In addition, data on climatic conditions, population density, population composition, and human travel patterns have also been collected. Many studies have employed the Johns Hopkins University CSSE database in conjunction with other databases to examine correlations between age and gender on the epidemic of COVID-19 (Hu et al., 2021), tourism and COVID-19 (Hu et al., 2022), weather, air pollution, and SARS-CoV-2 transmission (Xu et al., 2021), and the impact of heat waves (Lian et al., 2023).

Qualitative observation of the graphs generated by the Johns Hopkins University CSSE database revealed that death waves shared similar wave patterns in several countries. However, notable differences existed in various countries. For instance, a previous study examined whether there are similarities in the graphs of COVID-19 cumulative COVID-19 mortality rates between Canadian provinces and the American States (Myroniuk et al., 2023). Surprisingly, these authors found that most provinces and states are dissimilar in cumulative rates of COVID-19 mortality from January to December 2020 (Myroniuk et al., 2023), even the close ubication of the studied provinces. This led us to believe that a systematic similarity analysis of the worldwide number of COVID-19 deaths during the COVID-19 pandemic in the frequency domain could be helpful to characterize global COVID-19 mortality waves in detail. Therefore, the objectives of this study were: 1) to use the Johns Hopkins University CSSE database to characterize the power spectra of the worldwide COVID-19 mortality waves caused by COVID-19, and 2) to perform a similarity index analysis of the power spectra of the death waves among countries; this to quantitatively identify potential common patterns in the COVID-19 mortality waves.

## Results

We analyzed the daily COVID-19 death toll in 199 countries from January 22, 2020, to March 9, 2023, using data from the Johns Hopkins University CSSE database. Following an alphabetic order, we numbered the countries from 1 to 199, as illustrated in Table 1. We calculated the PSD of these time series per country. Figure 1A shows examples of time series from eight countries, whereas Figure 1B shows the respective PSD. Note that some countries exhibit a dominant peak around a frequency of one peak per year. After obtaining the PSD for all the countries listed in Table 1, we calculated the grand average of all PSD (n=199 countries), shown in Figure 2. In this global PSD, there are two dominant peaks of COVID-19 mortality waves, the first occurring at a frequency of 1.15 peaks per year and the second at 2.7 peaks per year.

**Figure 1.**
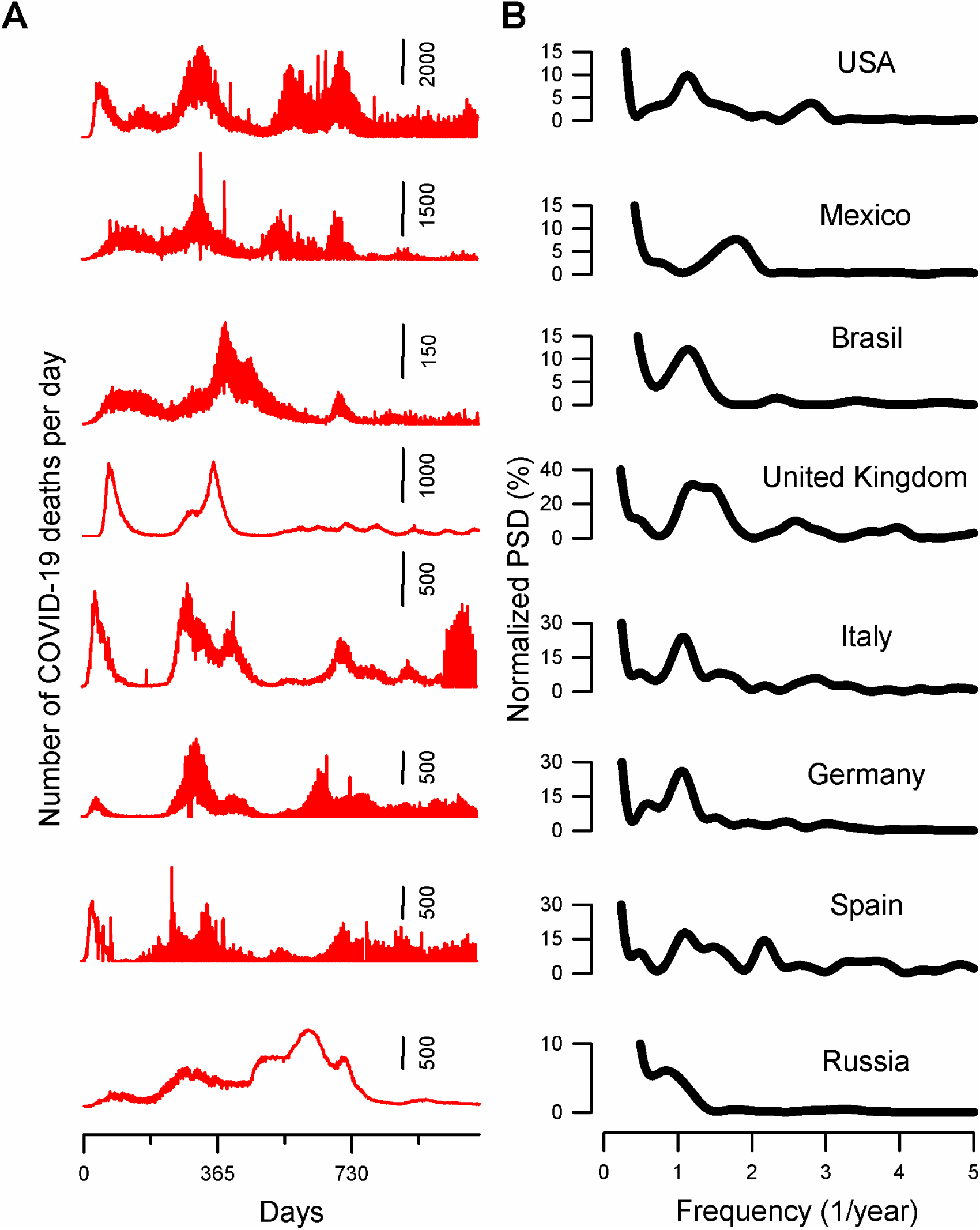
Examples of COVID-19 mortality waves and their power spectrum density (PSD). **A.** Examples of time series showing the number of COVID-19 deaths per day from eight countries. **B**. Normalized PSD is calculated from the time series shown in the left panel. Note that these countries exhibit a dominant peak around a frequency of one peak per year.

**Figure 2.**
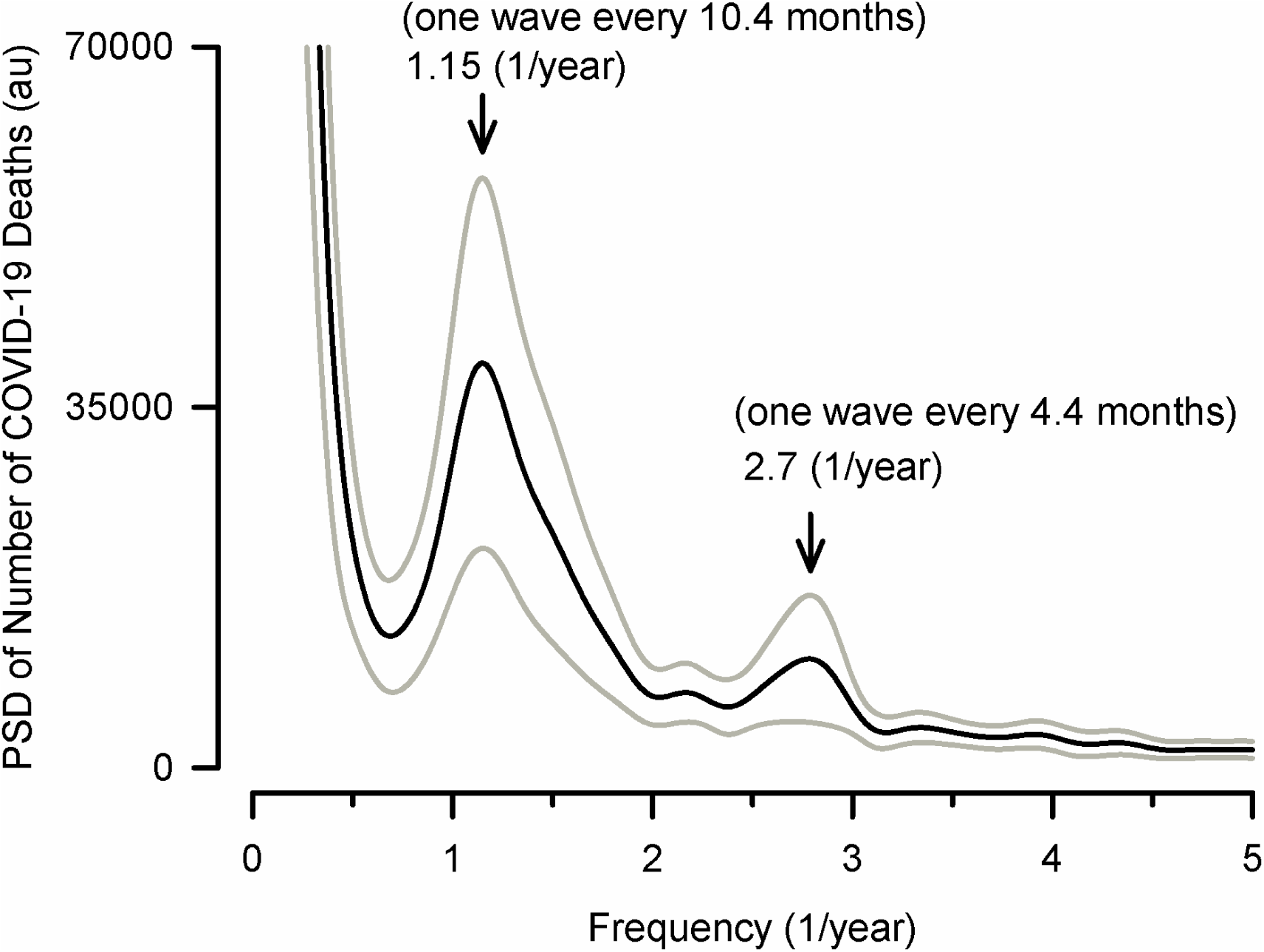
Global COVID-19 mortality waves worldwide. The blue trace is the grand average of the power spectrum density (PSD) obtained from the COVID-19 mortality time series for 199 countries. The traces in magenta color represent the standard deviation. Note the two dominant COVID-19 mortality waves occurring at frequencies of 1.15 waves/year (i.e., one wave every 10.4 months) and 2.7 waves/year (i.e., one wave every 4.4 months).

**Table 1.** A list of 199 countries was analyzed in the present study. Countries highlighted in green (n=17) correspond to those countries with PSD of low similarity in relation to all the countries. The other 182 countries not highlighted correspond to those countries with PSD of high similarity in relation to all the countries.

|  |  |  |  |  |
| --- | --- | --- | --- | --- |
| 1) Afghanistan | 41) Chile | 81) Guinea-Bissau | 121) Mauritius | 161) Saudi Arabia |
| 2) Albania | **42) China** | 82) Guyana | 122) Mexico | 162) Senegal |
| 3) Algeria | 43) Colombia | 83) Haiti | 123) Moldova | 163) Serbia |
| 4) Andorra | **44) Comoros** | 84) Honduras | 124) Monaco | 164) Seychelles |
| 5) Angola | 45) Congo | **85) Hong Kong** | 125) Mongolia | **165) Sierra Leone** |
| 6) Anguilla | 46) Costa Rica | 86) Hungary | 126) Montenegro | 166) Singapore |
| 7) Antigua and Barbuda | 47) Cote d'Ivoire | 87) Iceland | **127) Montserrat** | 167) Slovakia |
| 8) Argentina | 48) Croatia | 88) India | 128) Morocco | 168) Slovenia |
| 9) Armenia | 49) Curacao | 89) Indonesia | 129) Mozambique | 169) Somalia |
| 10) Aruba | 50) Cyprus | 90) Iran | 130) Myanmar | 170) South Africa |
| 11) Australia | 51) Czechia | 91) Iraq | 131) Namibia | 171) South Korea |
| 12) Austria | 52) Dem. Rep of Congo | 92) Ireland | 132) Nepal | 172) South Sudan |
| 13) Azerbaijan | 53) Denmark | 93) Isle of Man | 133) Netherlands | 173) Spain |
| 14) Bahamas | **54) Djibouti** | 94) Israel | 134) New Caledonia | 174) Sri Lanka |
| 15) Bahrain | 55) Dominica | 95) Italy | 135) New Zealand | 175) Sudan |
| 16) Bangladesh | 56) Dominican Republic | 96) Jamaica | 136) Nicaragua | 176) Sweden |
| 17) Barbados | 57) Ecuador | 97) Japan | 137) Niger | 177) Switzerland |
| 18) Belarus | 58) Egypt | 98) Jordan | 138) Nigeria | 178) Syria |
| 19) Belgium | 59) El Salvador | 99) Kazakhstan | 139) North Macedonia | 179) Taiwan |
| 20) Belize | 60) Equatorial Guinea | 100) Kenya | 140) Norway | 180) Tajikistan |
| 21) Benin | 61) Eritrea | **101) Kiribati** | 141) Oman | **181) Tanzania** |
| 22) Bermuda | 62) Estonia | 102) Kosovo | 142) Pakistan | 182) Thailand |
| 23) Bhutan | 63) Eswatini | 103) Kuwait | **143) Palau** | 183) Togo |
| 24) Bolivia | 64) Ethiopia | 104) Kyrgyzstan | 144) Palestine | 184) Trini. and Tobago |
| 25) Bonaire S. E. and Saba | **65) Faeroe Islands** | 105) Laos | 145) Panama | 185) Tunisia |
| 26) Bosnia and Herzegovina | 66) Fiji | 106) Latvia | 146) Papua New Guinea | 186) Turkey |
| 27) Botswana | 67) Finland | 107) Lebanon | 147) Paraguay | 187) Turks and C. Islands |
| 28) Brazil | 68) France | 108) Lesotho | 148) Peru | 188) Uganda |
| 29) British Virgin Islands | **69) French Polynesia** | 109) Liberia | 149) Philippines | 189) Ukraine |
| **30) Brunei** | 70) Gabon | 110) Libya | 150) Poland | 190) U. Arab Emirates |
| 31) Bulgaria | 71) Gambia | 111) Liechtenstein | 151) Portugal | 191) United Kingdom |
| 32) Burkina Faso | 72) Georgia | 112) Lithuania | 152) Qatar | 192) United States |
| **33) Burundi** | 73) Germany | 113) Luxembourg | 153) Romania | 193) Uruguay |
| 34) Cambodia | 74) Ghana | 114) Madagascar | 154) Russia | 194) Uzbekistan |
| 35) Cameroon | **75) Gibraltar** | 115) Malawi | 155) Rwanda | 195) Venezuela |
| 36) Canada | 76) Greece | 116) Malaysia | 156) Saint Kitts and Nevis | 196) Vietnam |
| 37) Cape Verde | **77) Greenland** | 117) Maldives | 157) Saint Lucia | 197) Yemen |
| 38) Cayman Islands | 78) Grenada | 118) Mali | 158) S. Vinc. and the Gre. | 198) Zambia |
| **39) Central African Rep.** | 79) Guatemala | 119) Malta | **159) San Marino** | 199) Zimbabwe |
| 40) Chad | 80) Guinea | 120) Mauritania | 160) Sao Tome and Princ. |  |

After examining the examples in Figure 1B, we noted some similarities in the PSD shape among these eight countries, which suggested that there would probably also be differences in the PSD shape among all 199 countries. Therefore, we computed the cosine similarity index between the PSD of all pairs of countries worldwide. The cosine similarity indexes obtained from PSD-shape comparisons are shown in the cosine similarity index matrix in Figure 3. The consecutive numbers in the horizontal and vertical axes represent the number assigned to each country according to Table 1. For clarity, we obtained the histogram of all cosine similarity indexes illustrated in Figure 3. Such a histogram is shown in Figure 4. It demonstrates the distribution of these cosine similarity indexes. The reader can observe that this distribution is negatively skewed. We calculated this distribution’s global cosine similarity index parameters, obtaining mean=0.8, median=0.84, mode=0.87, standard deviation=std=0.11, and skewness=-0.54.

**Figure 3.**
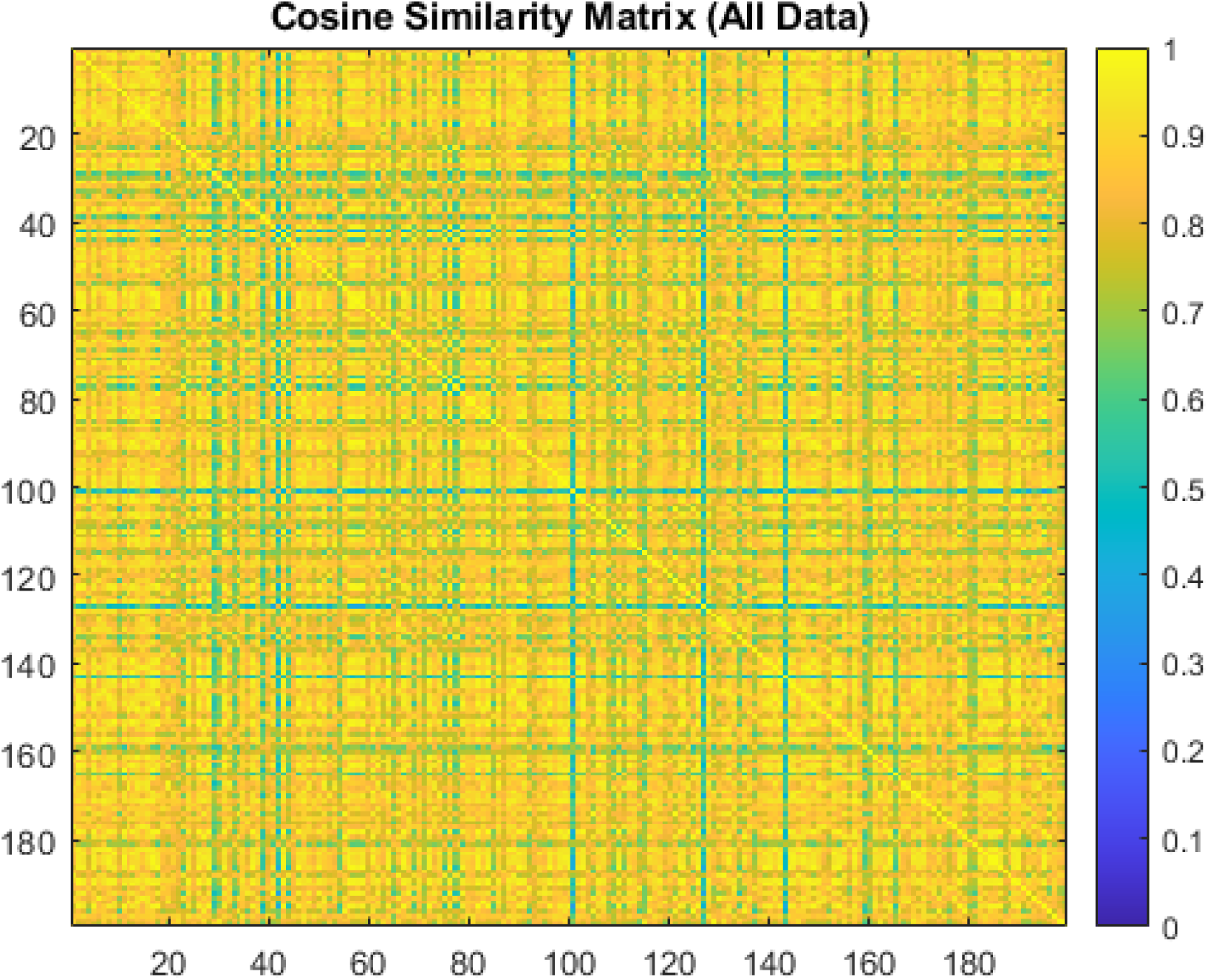
A cosine similarity matrix was obtained by comparing all pairs of COVID-19 mortality PSD for 199 countries listed in Table 1. The right vertical bar represents the cosine similarity index scale from 0 to 1.

**Figure 4.**
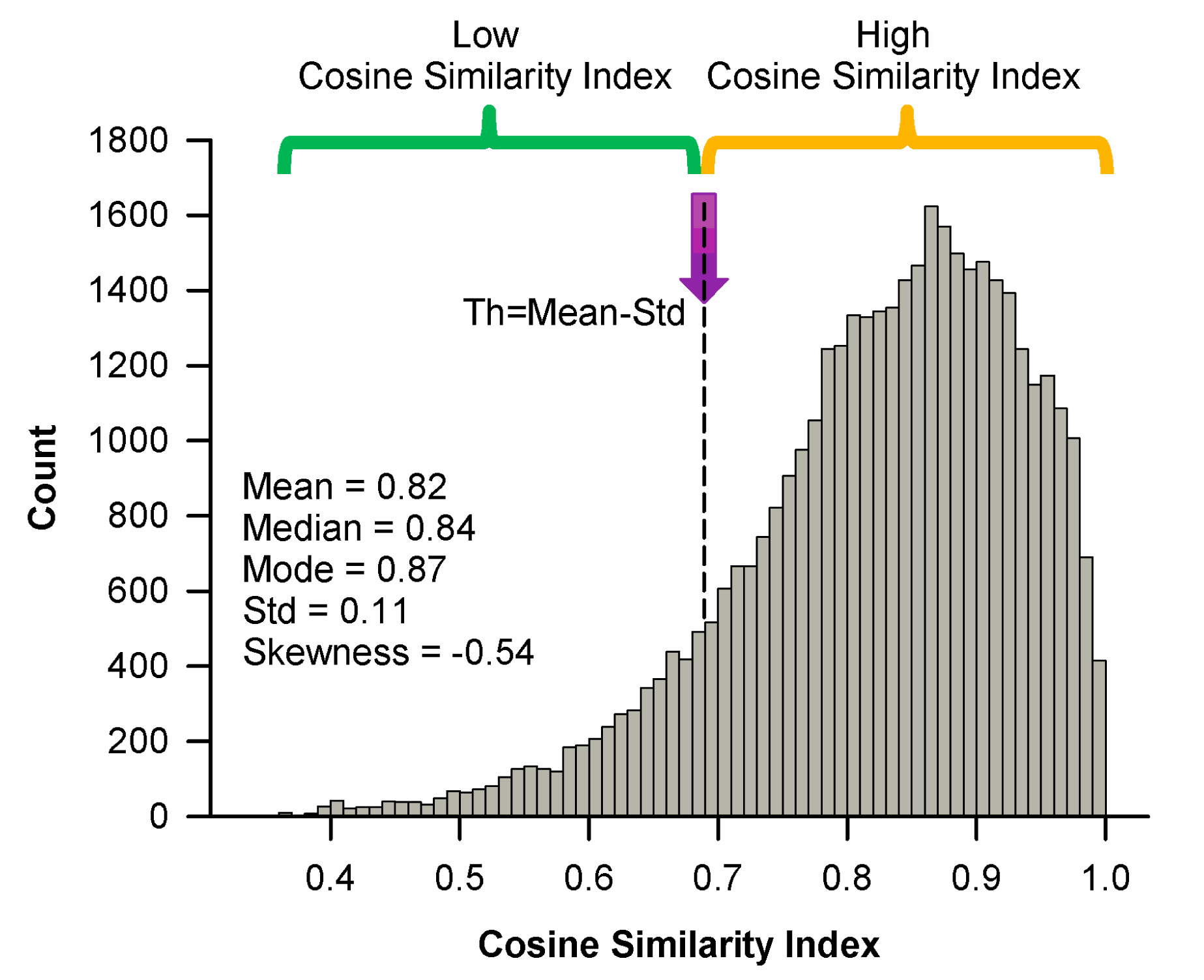
The histogram of the cosine similarity indexes was obtained from the cosine similarity index matrix shown in Figure 3. This histogram shows a negatively skewed distribution of the cosine similarity indexes for 199 countries with a skewness of −0.54. The formula Th=Mean-Std was used to calculate the threshold “Th” to separate a data group with low cosine similarity (green parenthesis) and a data group with high cosine similarity (yellow parenthesis). Std is for standard deviation.

After inspecting the colors in Figure 3, we can note some lines in the “green” color spectrum. These lines correspond to values below a cosine similarity index of 0.69, i.e., below the threshold Th=mean-std=0.8-0.11=0.69 of the negatively skewed distribution shown in Figure 4. We found that these “green” color spectrum lines correspond to 17 countries, as highlighted in green in Table 1. Some examples of the COVID-19 mortality time series and PSD obtained from these countries are illustrated in Figure 5. A notable qualitative characteristic of these COVID-19 mortality time series in Figure 5A is that the number of COVID-19 deaths per day does not exhibit COVID-19 mortality waves as those illustrated in Figure 1A. Another feature of these countries in Figure 5A is that they exhibited COVID-19 mortality waves in the PSD with multiple peaks (Figure 5B), thus exhibiting a different behavior in the COVID-19 mortality waves. For instance, China exhibited multiple dominant peaks and fewer COVID-19 deaths (see first panel of Figure 5B). Figure 6 is the grand average of the PSD obtained from these 17 countries with a cosine similarity index below 0.69. Note the absence of the two dominant peaks found in the global grand average of the PSD previously shown in Figure 2.

**Figure 5.**
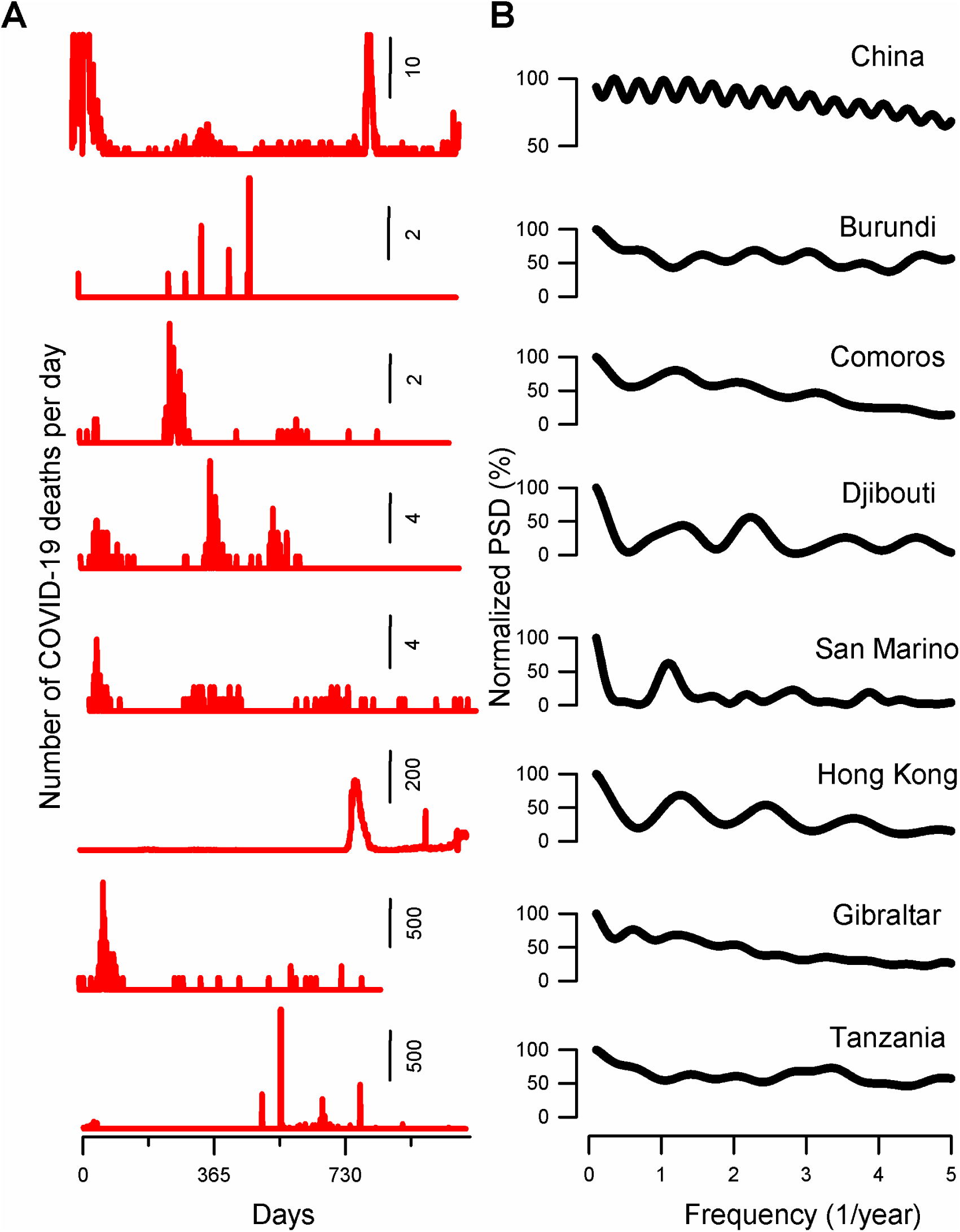
It has the same format as Figure 1 but for eight countries with a cosine similarity index below the threshold=mean-std=0.69. These countries exhibit a multi-peak behavior in their PSD, occurring at different frequencies.

**Figure 6.**
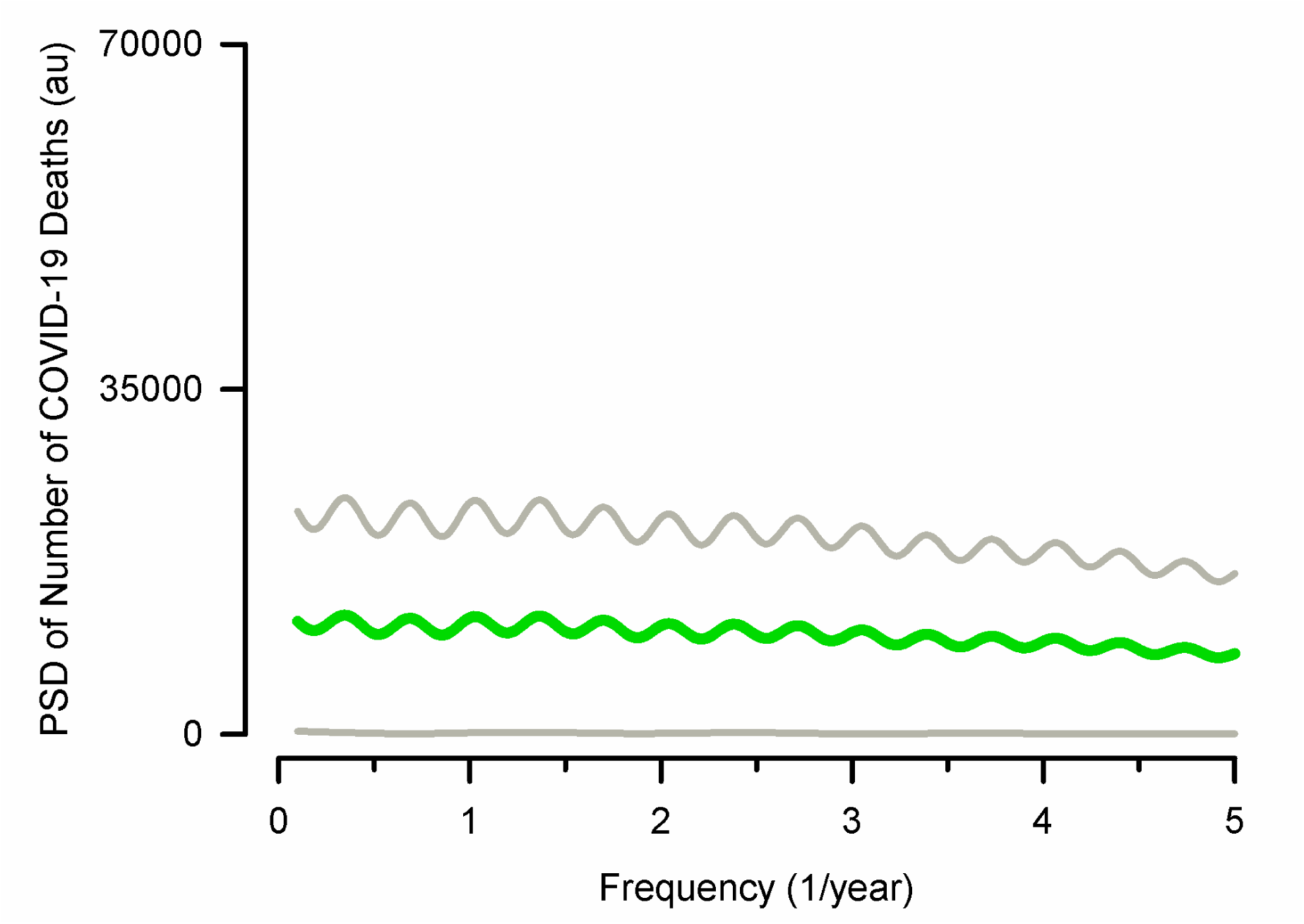
It has the same format as Figure 2, but the grand average of the PSD was obtained from 17 countries that exhibited a similarity index below 0.69. Note a multipeak behavior in the COVID-19 mortality waves occurring at different frequencies from 0.4 to 4.7 COVID-19 mortality waves per year.

Moreover, we found n=182 countries exhibiting a high cosine similarity index above the threshold Th=mean-std=0.69, which can be identified as those values in the “yellow” color spectrum in Figure 3. The grand average of the PSD for these 182 countries is shown in Figure 7.

**Figure 7.**
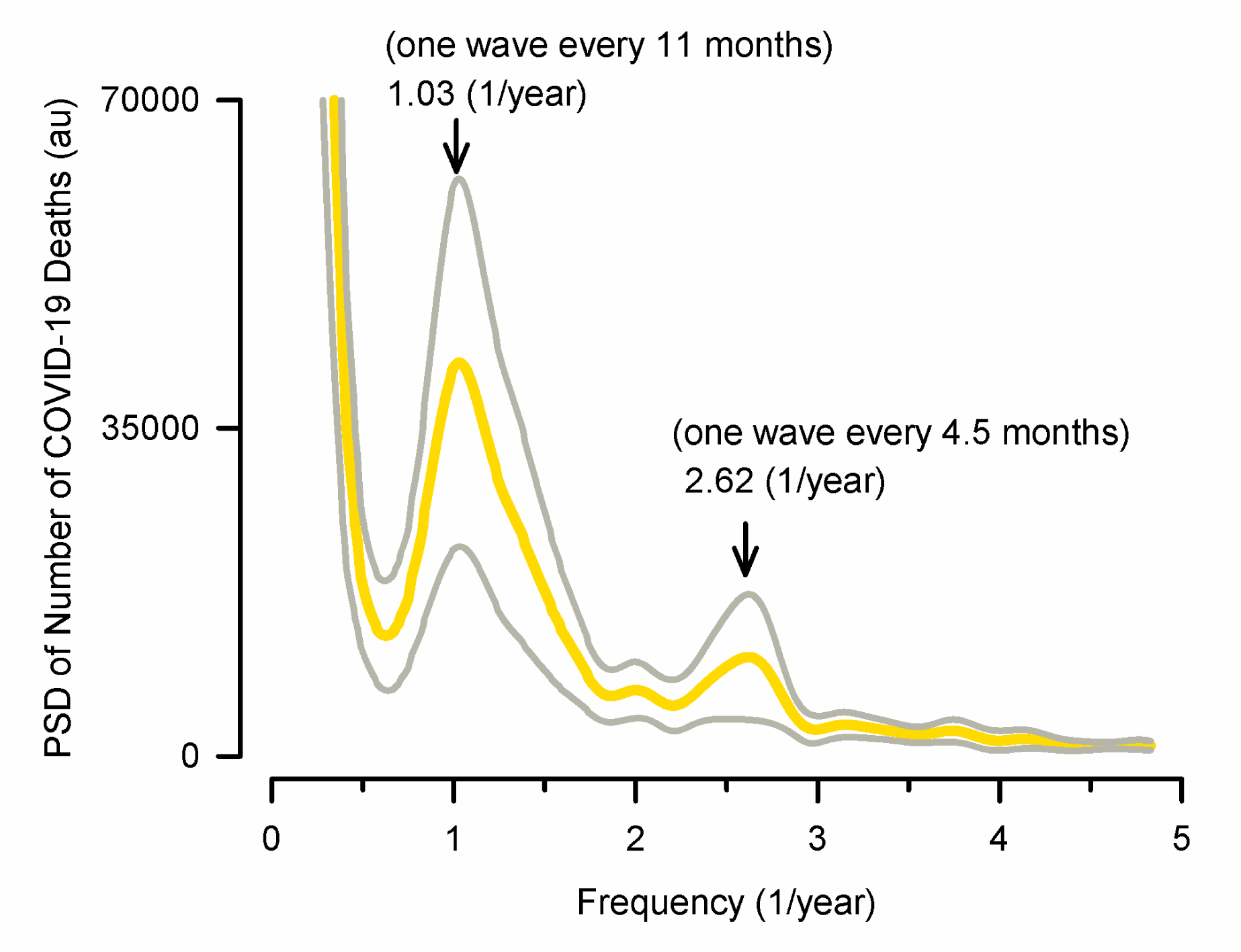
It has the same format as Figure 2 but for n=182 countries with a cosine similarity index above the threshold=mean-std=0.69. The averaged PSD for these 182 countries still exhibits two dominant peaks at similar frequencies of 1.03 and 2.62 peaks per year, as the grand averaged PSD.

Finally, we examined whether there is a statistically significant difference in the number of counts related to cosine similarity indexes below Th=0.69 versus those associated with cosine similarity indexes above Th=0.69. Both data groups are illustrated with parentheses in Figure 4. A non-parametric Mann-Whitney U test revealed a statistically significant difference p<0.0001 between both datasets. This result reveals that it helped define a cosine similarity index threshold of mean-std=0.69 as a criterion to justify the grand averaged PSD of countries with higher (Figure 7) and lower (Figure 6) cosine similarity indexes.

## Discussion

We analyzed the time series of COVID-19 mortality due to COVID-19 with a PSD. We identified two dominant peaks in the grand average of the PSD of 199 countries: one at a frequency of 1.15 waves per year (i.e., one wave every 10.4 months) and another at 2.7 waves per year (i.e., one wave every 4.4 months). After performing a cosine similarity analysis in the PSD shape for all countries, we found that most of these countries, n=182, exhibited a similar PSD shape with a high cosine similarity index above the threshold Th=mean-std=0.69, with two dominant peaks: one at 1.03 (i.e., one wave every 11 months) and another at 2.62 waves per year (i.e., one wave every 4.5 months). However, we found that n=17 countries exhibited a distinct PSD with a low cosine similarity index below the threshold Th=mean-std=0.69, with the feature that they exhibited a multipeak PSD shape.

We could speculate that the two COVID-19 mortality waves occurring at a periodicity of 11 months and 4.5 months may be correlated to the holiday periods, in which there is higher inter-country and intra-country human mobility. This suggestion is consistent with mathematical models predicting the impact of human mobility on the epidemic spread during holidays, highlighting that while inter-regional mobility could be a trigger for the epidemic spread, the diffusion effect of intra-regional mobility was primarily responsible for the outbreaks in a city (Li et al., 2023). In this context, it is possible that the differences in the COVID-19 mortality PSD waves among countries could be due to differences in governmental regulations, population density, weather conditions, and shared population activities without confinement, among other factors.

The observed similarities in the COVID-19 mortality PSD patterns among most countries validate the global nature of the pandemic, reflecting potential common factors influencing COVID-19 mortality rates (Figure 1 and Figure 3). However, the notable differences in PSD patterns among a subset of countries (Figure 6) versus another set of countries (Figure 7) highlight the potential influence of different local factors, such as public health policies, healthcare system resilience, and population behavior, in shaping the pandemic’s trajectory within specific contexts, among other unknown factors. Future studies will be necessary to uncover these factors. In this context, future research should identify other pandemic variables exhibiting a periodicity every 11 and 4.5 months.

We did a search in the literature for biological variables related to this periodicity of every 11 and 4.5 months, and we only found that infections in aquatic parasitic copepods (***Ch. Quaternia***) are increased with a periodicity of every 11 and 3 months in the oceanographic conditions of the Pacific Ocean associated with the 2015-2016 El Niño (Santana-Piñeros et al., 2020). The reader may note the parallelism in the periodicity of infection parameters in ***Ch. Quaternia*** and the periodicity of COVID-19 mortality due to SARS-CoV-2 infection in humans.

The findings from both the literature (Hass and Arsanjani, 2021; Dong et al., 2022; Hu et al., 2021; Hu et al., 2022; Xu et al., 2021; Lian et al., 2023) and the current study underscore the importance of global data collection and analysis tools, such as the Johns Hopkins University CSSE database, in monitoring and understanding pandemic trends. The variability in COVID-19 mortality patterns across countries emphasizes the need for tailored public health responses considering local conditions and capabilities. Furthermore, the identification of common COVID-19 mortality waves suggests potential areas for international collaboration in pandemic preparedness and response in case a future pandemic of a similar scale could occur. In this context, our study offers a characterization of the COVID-19 pandemic COVID-19 mortality waves, which could be helpful for future modeling studies and provide critical lessons for managing future global health crises.

A limitation of our study is the possibility that the counts of daily COVID-19 deaths are over- and under-estimated. Reports suggest that COVID-19 deaths were not counted adequately during the pandemic in several countries (Ioannidis, 2021). Another limitation is that we did not analyze correlations with potential factors that could influence the high and low cosine similarity in the periodicity of the COVID-19 mortality waves, such as governmental regulations, population density, weather conditions, and shared population activities without confinement, among other factors.

We conclude that the PSD analysis of COVID-19 mortality time series across 199 countries revealed patterns in COVID-19 mortality waves characterized by dominant peaks at frequencies of 1.15 and 2.7 waves per year. Moreover, the quantitative analysis using the cosine similarity index to compare the PSD shapes among countries uncovers a broad similarity in COVID-19 mortality patterns, with a significant portion of countries exhibiting high cosine similarity indices. However, it also identifies a subset of countries with distinct COVID-19 mortality wave patterns, as indicated by lower cosine similarity indices and multiple peaks in their PSDs. This divergence suggests variability in how different populations were impacted by and responded to the pandemic. Furthermore, the methodology employed here, using PSD analysis and the cosine similarity index, offers a novel approach to quantitatively assess global and local patterns of COVID-19 mortality, complementing previous qualitative observations that can be found in the Johns Hopkins University CSSE database (Dong et al., 2022).

## Materials and Methods

We employed the Johns Hopkins University CSSE COVID-19 database on GitHub.com to obtain the COVID-19 daily death counts for n=229 countries from January 22, 2020, to March 9, 2023.

### Inclusion and exclusion criteria

We analyzed daily COVID-19 death time series from 199 countries to obtain a power spectral density (PSD). We excluded 30 countries due to low COVID-19 deaths and the inability to obtain a PSD from their time series.

### Power Spectrum Density (PSD) and Cosine Similarity Index analysis

MATLAB was used to calculate the power spectrum density (PSD) of the time series of COVID-19 death counts in each country as follows:

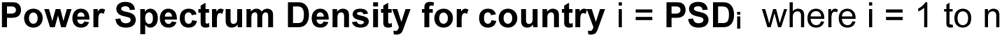

Because the PSD graphs have a characteristic shape that could be compared among countries, the PSD data for each country *i* was then used as a vector defined as:

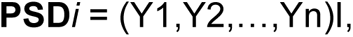

where Y1, Y2, .., Yn are the PSD values in the vertical axis of a PSD graph. Hence, we computed the cosine similarity index between all pairs of countries worldwide (n=199 countries) with these PSD vectors. This was the algorithm employed:

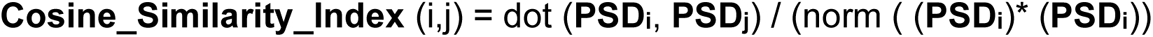

These cosine similarity indexes were plotted on a similarity matrix map to identify countries with notable differences in similarity index compared to the majority.

### Statistical analysis

Finally, we obtained a histogram of these cosine similarity indexes to identify the type of statistical distribution for the data. Then, skewness was calculated with the formula for Pearson’s second coefficient (Yule and Kendall, 1911) as follows:

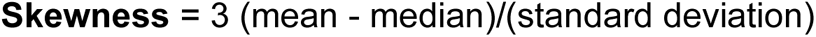

For statistical comparison, we separated two groups of cosine similarity index using a threshold “Th” defined by “Th=mean-std,” where “std” is the standard deviation. This threshold helped identify countries with a low cosine similarity index below “mean-std” and countries with a high cosine similarity index above “mean-std.” Because data were not normally distributed, we employed a non-parametric Mann-Whitney U test to compare the incidence between these two groups of cosine similarity indexes.

## Data Availability

All data produced in the present study are available upon reasonable request to the authors

